# Beta-blockers Increase Cardiovascular Mortality in Hemodialysis Patients with Orthostatic Hypotension

**DOI:** 10.1101/2024.09.30.24314645

**Authors:** M. Schiavone, C Castellaro, JC. Pereira Redondo, C. Diaz, G. Laham

**Author notes:** **Corresponding author** Maximo A. Schiavone, MD., Centro de Educación Médica e Investigaciones Clínicas Norberto Quirno (CEMIC), Galvan 4102, Ciudad Autónoma de Buenos Aires, Argentina.

## Abstract

**Background:** Orthostatic hypotension (OH) is prevalent among dialysis patients and is a known cardiovascular (CV) risk factor. Beta-blockers (BBs) are commonly prescribed to mitigate CV mortality in this population, despite potential risks associated with OH. This study examines the impact of BBs on CV mortality among dialysis patients with OH.

**Methods:** We conducted a prospective analysis of 134 dialysis patients from the PRECADIA program at the Centro de Educación Médica e Investigaciones Clínicas (CEMIC), focusing on hemodynamic assessments including blood pressure changes from supine to standing positions, and evaluating CV mortality over a 3-year follow-up. OH was defined by a decrease of ≥10 mmHg in diastolic blood pressure or ≥20 mmHg in systolic blood pressure upon standing. Cox regression analyses were utilized to identify independent predictors of CV mortality.

**Results:** Of the patients, 23.1% were identified with OH. No significant differences in demographic or baseline clinical characteristics were observed between patients with and without OH, except for a higher diabetes prevalence in the OH group. OH patients treated with BBs demonstrated significantly higher CV mortality (29.6%) compared to those not receiving BBs. Independent predictors of CV mortality included age, time on dialysis, and BB use, with BBs significantly associated with increased CV mortality risk.

**Conclusions:** Dialysis patients with OH exhibit a high CV mortality rate, significantly influenced by BB usage. While BBs are recommended for managing CV risks in dialysis patients, their association with increased CV mortality in patients with OH necessitates careful consideration and management strategies.

## INTRODUCTION

Orthostatic hypotension (OH) is characterized by a significant decrease in an individual’s blood pressure (BP) when changing from a supine to standing position. Clinically, a reduction of ≥ 20 mmHg in systolic blood pressure (SBP) and/or ≥ 10 mmHg in diastolic blood pressure (DBP) within the first three minutes of standing upright is defined as OH [1, 2]. OH is a well-known predictor of mortality and morbidity due to cardiovascular (CV) causes in the general population [4-7]. In the recently conducted SPRINT (Systolic Blood Pressure Intervention Trial), Townsend et al. [8] showed a significant association between the prevalence of OH and cardiovascular diseases, including heart failure, peripheral arterial disease, and atrial fibrillation among the general population. In 2010, Shibao and Biaggioni [9] also reported that OH significantly increased the risk of heart attack, stroke, heart failure, and mortality among the general population. In a recent meta-analysis by Saedon et al. [3], the investigators reported an overall OH prevalence of 23.9% (95% CI: 18.2-30.1) among the general population. In addition, Januszko-Giergielewicz et al. [10] reported an OH prevalence of 37.8% among chronic kidney disease (CKD) patients.

Beta-blockers (BBs) are widely used for OH management. These are competitive antagonists that counteract the sympathetic action of catecholamines on beta-adrenergic receptors [11]. BBs are also recommended for dialysis patients to reduce mortality [12]. However, some studies have shown a correlation between the use of BBs and OH development in CKD patients. For instance, Januszko- Giergielewicz et al. [10] reported a significant association between the use of BBs and OH prevalence (p < 0.04) for an estimated glomerular filtration rate range of 30-60 mL/min/1.73 m^2^.

There have been limited studies on the prevalence of OH among dialysis patients [11, 13-15]., Panuccio et al. [17] assessed the correlation between OH incidence and the survival of individuals on peritoneal dialysis (PD). They reported an OH prevalence of 30% among PD patients, of which 18% had systolic and 12% had diastolic OH. They also reported that 11% of PD patients exhibited CV mortality.

In the present study, we analysed the association between OH and the risk of CV mortality and whether BB can increase the risk of mortality among dialysis patients.

## MATERIALS AND METHODS

### Population

Initially, 134 patients were included in this prospective study from a CV programme that assessed patients on dialysis (PRECADIA) at the Centro de Educación Médica e Investigaciones Clínicas (CEMIC). The patients were observed in a structured and standardized manner during inter-dialysis days. Data were collected with respect to weight, height, standardized brachial BP, transthoracic echocardiography, and non-invasive haemodynamic assessment via impedance cardiography (ICG). Patients with an incomplete haemodynamic analysis, amputation, or reduced ejection fraction (below 40%) were excluded from the analysis.

Dialysis patients were subsequently divided into NoOH and OH groups based on the absence or presence of OH, respectively. OH was defined as a decrease of ≥10 mmHg in DBP or ≥ 20 mmHg in SBP when assuming a standing position.

### Haemodynamic assessment

Brachial BP was determined in the fistula-free arm, following the Argentine Society of Hypertension protocol, using Mobil O Graph equipment (I.E.M. GmbH, Stolberg, Germany) with the patient in the sitting position. The patient was placed in the supine position for three minutes during the procedure. BP was then measured in the same position. Similarly, the patients were then assessed haemodynamically but in the standing position for three minutes. At the end of this period, a final BP was measured.

### Analysed variables

In the current study, age, sex, body mass index (BMI), time on dialysis, anti-hypertensive treatment, and a history of CV events (myocardial infarction and/or stroke) were recorded. The following haemodynamic variables were analysed: SBP, DBP, in the supine and standing positions after three minutes.

### Primary endpoint

After the CV assessments, a 3-year follow-up was conducted to evaluate the cause of any mortality that occurred. We defined CV mortality as patient demise attributed to sudden death, acute myocardial infarction, and/or cardiogenic shock.

### Statistical analysis

Haemodynamic variables were analysed according to basal conditions and the groups’ differences (∆ standing − lying). Data were expressed as either medians and interquartile ranges or means and standard deviations. Differences between the continuous and categorical variables were determined using independent samples t-tests and chi-square tests, respectively. Survival between both groups was compared using log-rank and Kaplan-Meier tests.

Cox regression analyses were used to determine independent mortality predictors, such as age, gender, BP, time on Dx, diabetes diagnosis, treatment with BBs, and OH. A second cox regression analysis was performed to compare the CV mortality of OH patients with and without treatment with BBs. This analysis was conducted after adjusting for age, gender, time on dialysis, and diagnosis of diabetes.

Med Calc, version 13, was used for statistical analysis. Statistical significance was defined as p <0.05.

## RESULTS

### Patient recruitment and demographic data

Initially, 134 patients were enrolled in the study. Based on the exclusion criteria, 17 patients were later excluded. The overall mean patient age was 59.72 ± 16 years. Approximately 44.4% of the patients were female. The mean SBP and DBP of the patients were 147.4 ± 31 and 86.75 ± 17 mmHg, respectively. Of the 117 patients in the study, 27 (23.1%) presented with OH, while 90 were grouped into the NoOH group. Differences in age, sex, BMI, dialysis vintage, anti-hypertensive treatment, and CV events were not significant between the groups (Table 1). However, patients in the OH group exhibited a higher prevalence of diabetes (p = 0.003).

**Table 1.**
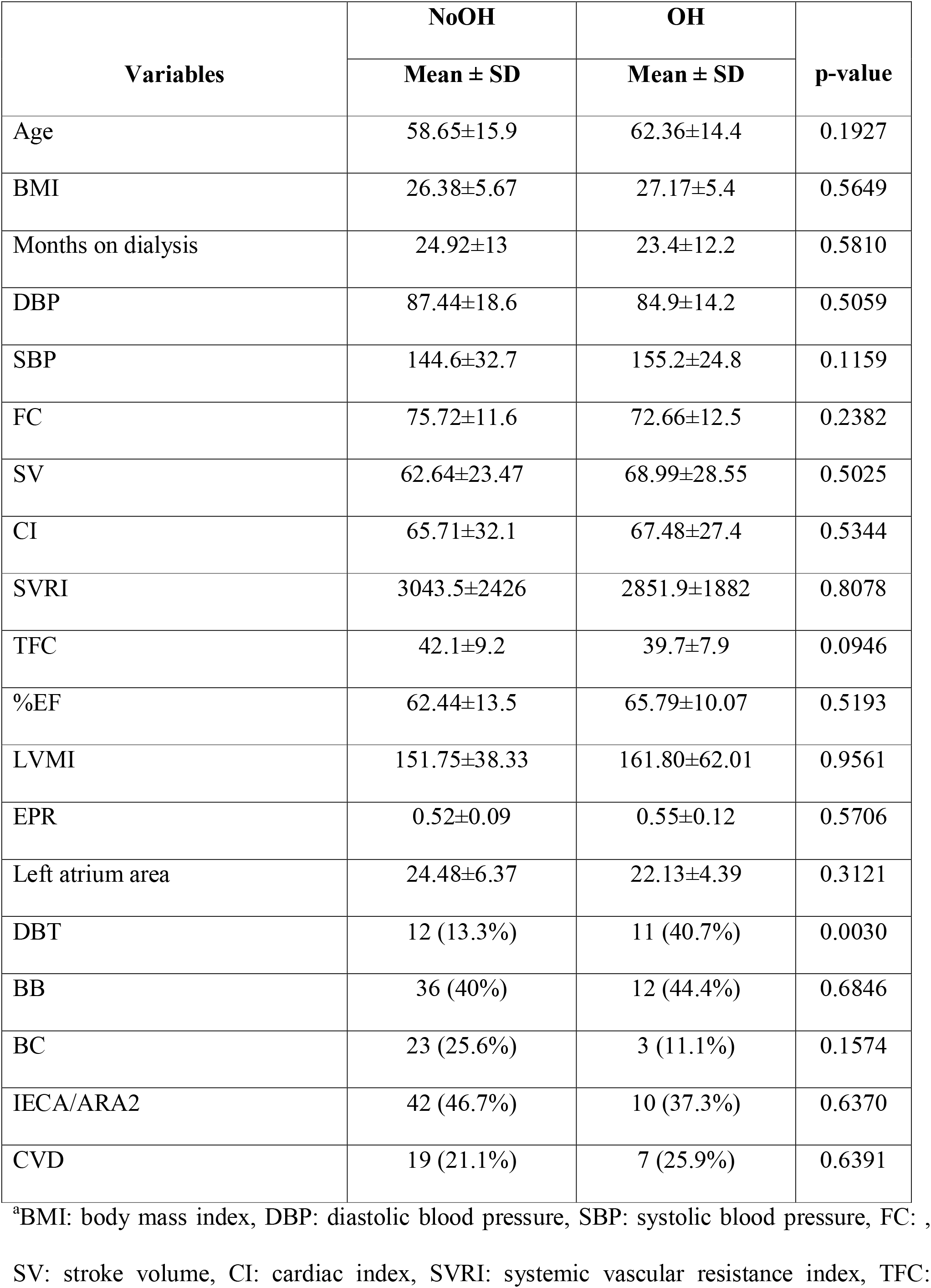
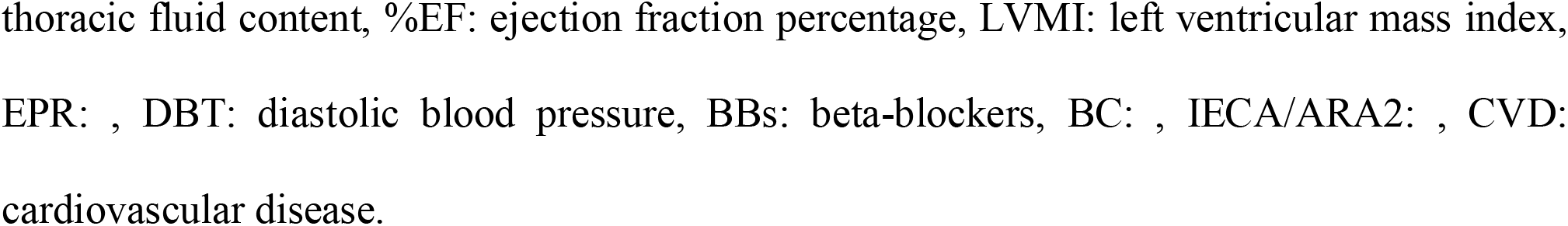
Baseline data of the study population.

| Variables | NoOH | OH | p-value |
| --- | --- | --- | --- |
| | Mean ± SD | Mean ± SD | |
| Age | 58.65±15.9 | 62.36±14.4 | 0.1927 |
| BMI | 26.38±5.67 | 27.17±5.4 | 0.5649 |
| Months on dialysis | 24.92±13 | 23.4±12.2 | 0.5810 |
| DBP | 87.44±18.6 | 84.9±14.2 | 0.5059 |
| SBP | 144.6±32.7 | 155.2±24.8 | 0.1159 |
| FC | 75.72±11.6 | 72.66±12.5 | 0.2382 |
| SV | 62.64±23.47 | 68.99±28.55 | 0.5025 |
| CI | 65.71±32.1 | 67.48±27.4 | 0.5344 |
| SVRI | 3043.5±2426 | 2851.9±1882 | 0.8078 |
| TFC | 42.1±9.2 | 39.7±7.9 | 0.0946 |
| %EF | 62.44±13.5 | 65.79±10.07 | 0.5193 |
| LVMI | 151.75±38.33 | 161.80±62.01 | 0.9561 |
| EPR | 0.52±0.09 | 0.55±0.12 | 0.5706 |
| Left atrium area | 24.48±6.37 | 22.13±4.39 | 0.3121 |
| DBT | 12 (13.3%) | 11 (40.7%) | 0.0030 |
| BB | 36 (40%) | 12 (44.4%) | 0.6846 |
| BC | 23 (25.6%) | 3 (11.1%) | 0.1574 |
| IECA/ARA2 | 42 (46.7%) | 10 (37.3%) | 0.6370 |
| CVD | 19 (21.1%) | 7 (25.9%) | 0.6391 |

<sup>a</sup>BMI: body mass index, DBP: diastolic blood pressure, SBP: systolic blood pressure, FC: , SV: stroke volume, CI: cardiac index, SVRI: systemic vascular resistance index, TFC:
thoracic fluid content, %EF: ejection fraction percentage, LVMI: left ventricular mass index, EPR: , DBT: diastolic blood pressure, BBs: beta-blockers, BC: , IECA/ARA2: , CVD: cardiovascular disease.

### Haemodynamic assessment

Differences between the haemodynamic variables of both groups were not significant. When assuming the standing position, patients in the OH group showed significantly higher ∆DBP and ∆SBP than those in the NoOH group (∆DBP and ∆SBP; NoOH: 3.76 ± 8.1 mmHg and 3.53 ± 15.8 mmHg vs OH: -11.6 ± 8.5 mmHg and -32.28 ± 18 mmHg; both p < 0.0001).

### Survival during follow-up

The median follow-up period was 36.2 months (95% CI: 34.1-38.3), during which 17 patients (14.5%) died of CV causes–eight of whom belonged to the OH group. The patients in the OH group exhibited a significantly higher mortality rate than those in the NoOH group (29.6% and 10%, respectively, p = 0.02). In addition, patients in the NoOH group exhibited a higher mean survival duration of 37.8 months (range: 35.8-39.8 months) compared to the OH patients, with a mean survival duration of 32.08 months (range: 26.9-37.3 months). The differences between the survival probabilities of both groups are illustrated in Figure 1.

**Figure 1:**
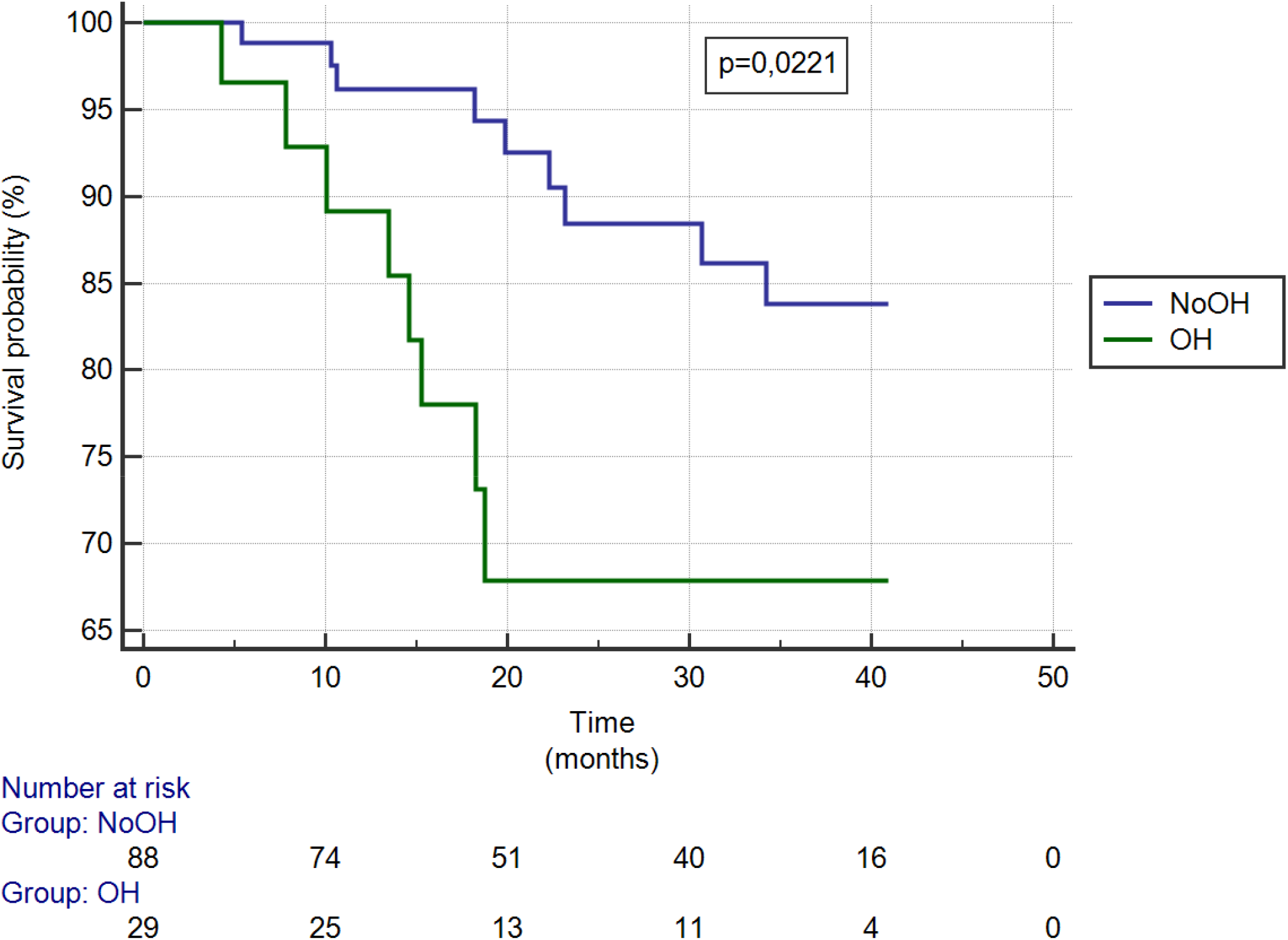
The differences between the survival probabilities of the OH and NoOH groups

### Regression analysis

Tables 2 and 3 depict the results of the Cox proportional hazard regression (HR) analyses. As can be seen from Table 2, without any adjustment, among all of the analysed factors, age (HR: 1.06, 95% CI: 1.02-1.1, p = 0.0019), time on dialysis (HR: 1.00, 95% CI: 1.00-1.01, p = 0.0313), presence of OH (HR: 2.9, 95% CI: 1.12-7.53, p = 0.0289), diabetes (HR: 3.44, 95% CI: 1.3-9.1, p = 0.0128), and use of BBs (HR: 6.96, 95% CI: 1.98-28.48, p = 0.0025) were significantly correlated with CV mortality. After adjusting for confounders, age (HR: 1.07, 95% CI: 1.02-1.11, p = 0.0032), time on dialysis (HR: 1.01, 95% CI: 1.00-1.01, p = 0.0395), and use of BBs (HR: 10.04, 95% CI: 2.45-41.2, p = 0.0014) were found to be independently associated with CV mortality (Table 2).

**Table 2.**
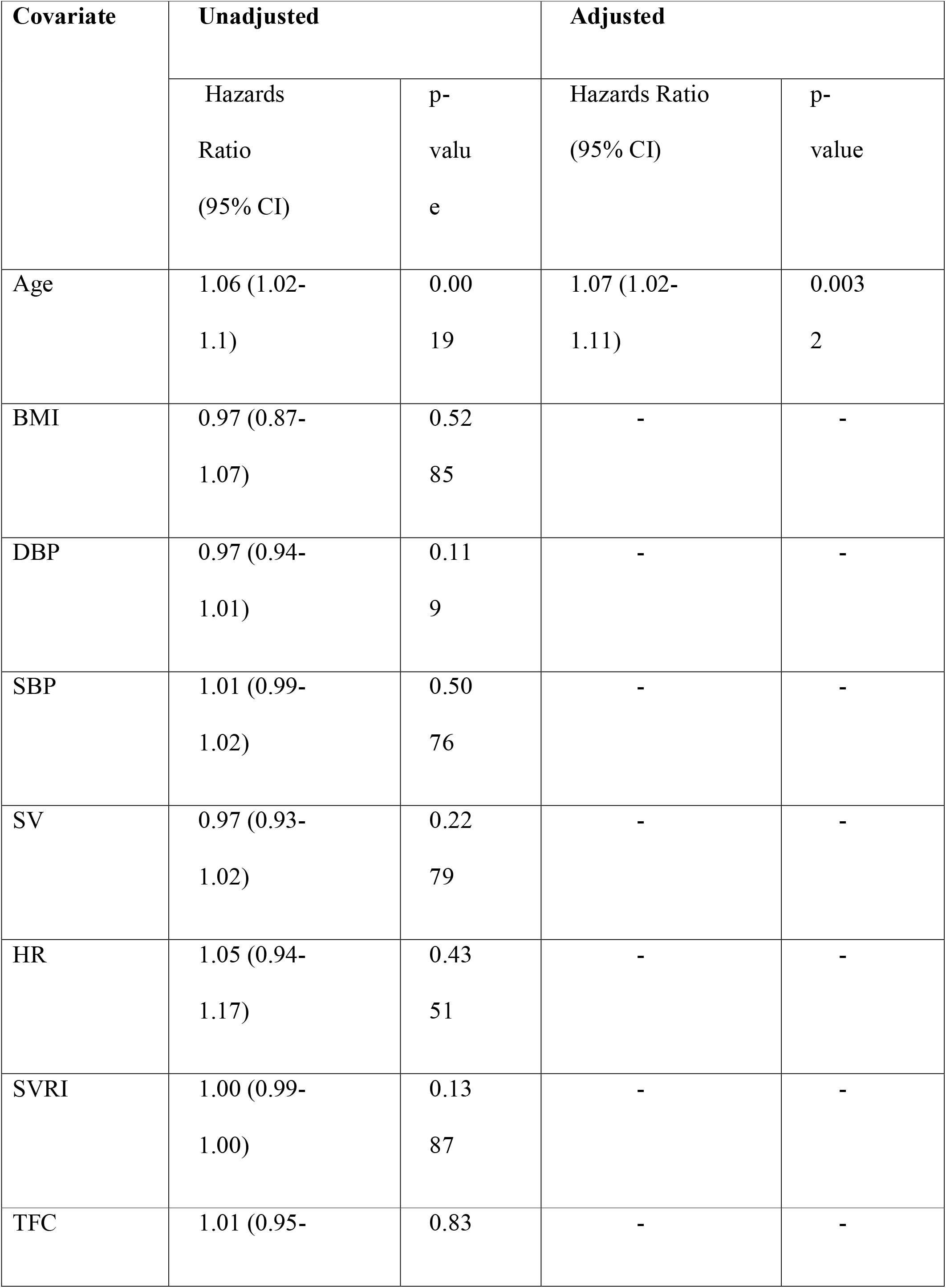
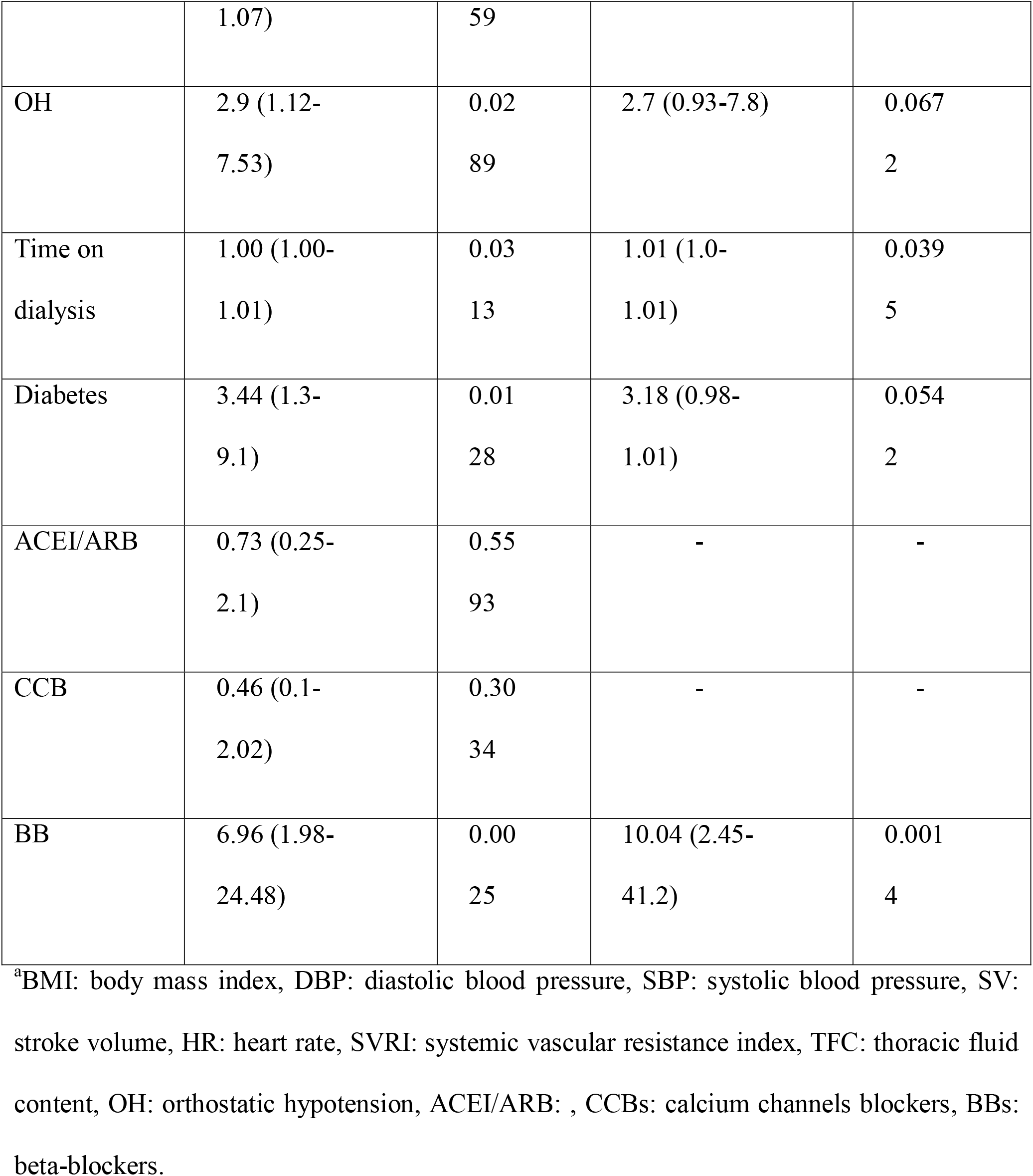
Univariate and multivariate Cox regression analysis for factors associated with cardiovascular mortality in dialysis patients.

**Table 3.** Univariate and multivariate Cox regression analyses comparing cardiovascular mortality among patients with and without OH (and either treated of not or with BBs).

| Covariate | Unadjusted |  | Adjusted* |  |
| --- | --- | --- | --- | --- |
|  | HR<br>(95%<br>CI) | p-value | HR<br>(95%CI) | p-value |
| NoOH | Ref. | - | Ref. | - |
| OH<br>without<br>t BB | 1.23<br>(0.27-5.68) | 0.7<br>94<br>7 | 0.39<br>(0.07-2.2) | 0.2<br>86<br>1 |
| OH<br>with<br>BB | 5.33<br>(1.9-15.04<br>) | 0.0<br>01<br>6 | 5.48<br>(1.79-16.77) | 0.0<br>02<br>9 |

\*adjusted for age, gender, time on dialysis and diagnosis of diabetes.
<sup>a</sup>HR: hazards ratio, OH: orthostatic hypotension, BBs: beta-blockers.

The patients in the OH group were further divided into two subgroups based on whether they were treated with BBs. As shown in Table 3, both before and after adjusting for confounders, the OH patients treated with BBs experienced significantly higher CV mortality (before and after adjustment: p < 0.0016 and p < 0.0029, respectively) than OH patients who did not receive them. Patients with OH that do not receive BB showed no difference in mortality when compared to

NoOH patients. The differences between the survival probabilities of the three groups are illustrated in Figure 2.

**Figure 2:**
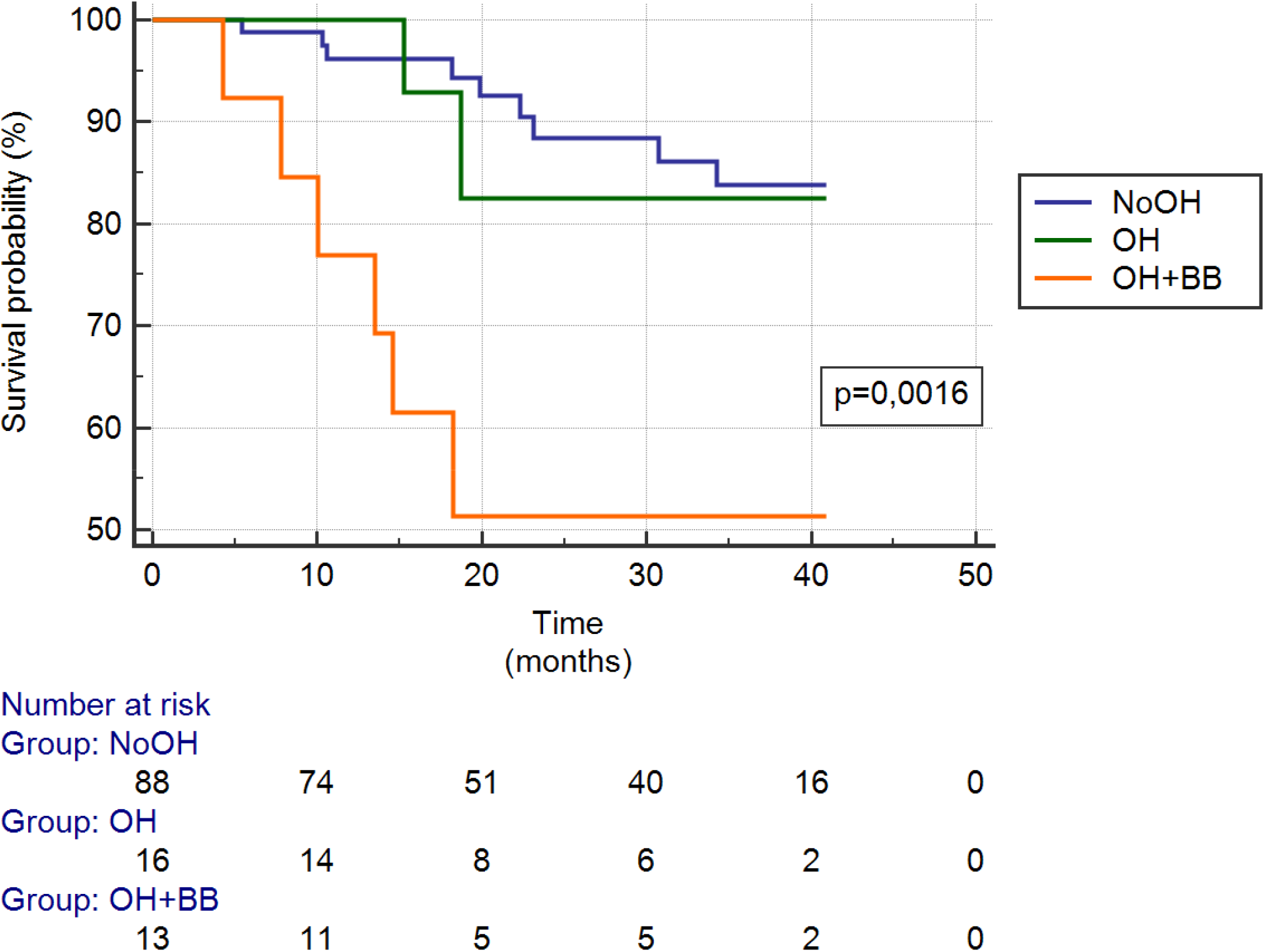
The differences between the survival probabilities of the OH with BB, OH without BB and NoOH groups

## DISCUSSION

In this study, we observed a high prevalence of OH in our sample.

The use of BBs appeared to affect the risk of CV mortality. In the Cox regression analyses, the use of BBs was identified as an independent predictor of CV mortality in OH patients (Table 3). Previous studies have shown that BBs are widely used in CKD patients due to the elevated risk of CV disorders and the proven therapeutic effect of BBs on CV morbidity and mortality [19-21]. In addition, BBs have previously been recommended in dialysis patients to alleviate arterial hypertension [22]. This subgroup of patients has exhibited sympathetic hyperactivity, which, in turn, is a potent predictor of CV death [23]. According to the United States Renal Data System (USRDS), dialsysis patient’s primary causes of death includes severe arrhythmia and sudden death, thereby rendering a protective role for BBs [24, 25]. The Dialysis Outcomes and Practice Patterns Study also demonstrated that BB administration to Dx patients is associated with a low risk of sudden death [26].

There is a high prevalence of OH among patients on Dx. This complication arises due to autonomic dysfunction. It is also responsible for patients’ poor quality of life and high morbidity and mortality due to CV disorders. Previous studies have described a dysautonomia incidence of 60-80% among Dx patients and the use of midodrine to treat this complication [27-30]. In this study, we observed an OH prevalence of 23.1% among our study group. This was slightly lower than that reported by Panuccio et al. [17], who found an OH prevalence of 30% among Dx patients. The lower OH prevalence in our sample could be attributed to the low number of recruited patients and the lower overall mean age of our patients, as it has been shown that OH prevalence increases with age.

The 3-year follow-up of our study population revealed significantly higher CV mortality among the patients in the OH group than in the NoOH group. Furthermore, we observed a higher risk of CV mortality among OH patients who received BBs compared to those who did not, which suggests that the use of BBs did not exert any protective effects against CV mortality in our patients. It is noteworthy that our patients also carried comorbidities. However, we observed that CV mortality was only significantly associated with the presence of diabetes mellitus. Our results agree with those by Hateren et al. [31], who reported a higher OH prevalence in diabetes patients and showed that the presence of diabetes was an independent predictor of OH. In another recent study, Parimala and Surendhar [32] also reported a high prevalence of OH among diabetes patients, with a majority exhibiting OH in the standing position. In this study, none of the other comorbidities investigated was found to be significantly associated with a higher CV mortality of OH patients treated with BBs (Supplementary Table 1).

There were a few limitations of this study. First, we recruited a relatively low number of patients, and the number of patients with OH was even less. This might affect the generalizability of our results. Second, in the Cox regression analyses (and after adjusting for confounders), although the presence of OH and diabetes appeared to independently affect the risk for CV mortality, the correlation was just barely significant. This result could be refined by increasing the sample size.

The risk of CV mortality among Dx patients with OH was significantly correlated with using BBs, which did not impart any significant protective effects against CV disorders in our patients. The patients in the OH group who were treated with BBs exhibited the lowest probability of survival, followed by those in the OH group without BB treatment, and finally those in the NoOH group.

## Data Availability

All data produced in the present work are contained in the manuscript

## ACKNOWLEDGEMENTS

Maria Elena Biain MD, Carlos Callegari MD and Mauro Magenta MD.

